# Deletion size and background genetic variation shape congenital heart disease phenotypes in 3,016 individuals with 22q11.2 deletion syndrome

**DOI:** 10.64898/2026.02.23.26346918

**Authors:** Jhih-Rong Lin, Daniella Miller, Dana Luong, Tanner Nelson, T. Blaine Crowley, Oanh T. Tran, Bhooma Thiruvahindrapuram, Amirhossein Hajianpour, Linda Campbell, Tiffany Busa, Damian Heine-Suñer, Sixto García-Miñaúr, Luis Fernández, Kieran C. Murphy, Declan Murphy, Wanda Hawula, Kathleen Angkustsiri, Vandana Shashi, Kelly Schoch, Carrie E. Bearden, Aoy Tomita Mitchell, Michael E. Mitchell, Miri Carmel, Abraham Weizman, Elena Michaelovsky, Doron Gothelf, Marianne B.M. van den Bree, Michael J. Owen, Jacob A.S. Vorstman, Erik Boot, Claudia Vingerhoets, Therese van Amelsvoort, Ann Swillen, Jeroen Breckpot, Joris R. Vermeesch, Koen Devriendt, Maude Schneider, Stephan Eliez, M. Cristina Digilio, Marta Unolt, Carolina Putotto, Paolo Versacci, Bruno Marino, Maria Pontillo, Marco Armando, Stefano Vicari, Gabriela M. Repetto, Wendy R. Kates, Robert J. Shprintzen, Raquel E. Gur, Elaine H. Zackai, Elizabeth Goldmuntz, Tao Wang, Srilakshmi Raj, Beverly S. Emanuel, Donna M. McDonald-McGinn, Stephen C. Scherer, Anne S. Bassett, Zhengdong D. Zhang, Bernice E. Morrow

## Abstract

Congenital heart disease (CHD) occurs in over half of individuals with 22q11.2 deletion syndrome (22q11.2DS), but lesion type varies widely. We analyzed 3,016 unrelated postnatal individuals with 22q11.2DS from specialized centers in the United States, Canada, Europe, South America, Israel, and Australia, including 1,868 with whole-genome sequencing. The typical 3 Mb LCR22 A-D deletion was present in 2,788 individuals (92.4%), with smaller A-B (n=172, 5.7%) and A-C (n=56, 1.9%) deletions comprising the remaining cohort. In multivariable mixed-effects logistic regression adjusting for sex, deletion group, genome-wide principal components (PCs), and recruitment site, four non-intercept associations, to test relationships, met study-wide FDR statistical significance. Compared with the A-B deletion, the A-D deletion was associated with lower odds of persistent truncus arteriosus (OR=0.37, 95% CI 0.18-0.75) and higher odds of isolated septal defects (OR=4.68, 95% CI 1.71-12.83), although precision was limited by the smaller A-B group. PC2 was associated with lower odds of pulmonary stenosis/atresia with additional lesions (OR=0.73, 95% CI 0.61-0.87), and PC4 with higher odds of abnormal origin of the subclavian arteries (OR=2.59, 95% CI 1.37-4.89). ADMIXTURE-derived continental ancestry proportions did not show independent associations with these two PC-associated outcomes. These lesion-specific findings suggest the hypothesis that deletion interval and broader genetic background may contribute to CHD variability in 22q11.2DS, pending future replication.

**KEY MESSAGES:** *What is already known on this topic:* Chromosome 22q11.2DS is a rare genetic disorder associated with serious medical challenges. Most affected individuals have congenital heart disease but with variable phenotypic expression. One of the main questions still an open topic in the field is why individuals with 22q11.2DS show extensive phenotypic heterogeneity.

*What this study adds:* Analysis of retrospective data on 3,016 individuals with 22q11.2DS and 1,868 with sequence data suggest that deletion type and genetic variation influence phenotypic expression of congenital heart disease.

*How this study might affect research, practice or policy:* This work informs the clinician as to the importance of testing for deletion type and obtaining accurate cardiac phenotypes to diagnose 22q11.2DS that will improve the prognosis and disease progression. It also implicates the complexity of sex and ancestry as confounding factors that will help guide future research studies to identify genetic modifiers of congenital heart disease.

## INTRODUCTION

The chromosome 22q11.2 deletion syndrome (22q11.2DS; DiGeorge/velo-cardio-facial/conotruncal anomaly face syndrome) is a congenital anomaly disorder that occurs in 1/1,000 fetuses [1];[2] and 1/2,150 live births [3–5] constituting the most frequent genomic disorder in humans. This syndrome results from non-allelic homologous recombination events between flanking low copy repeats on chromosome 22q11.2 termed LCR22 [6–9] resulting in *de novo* deletions of different sizes (**Figure 1**). Molecular genetic testing such as multiplex-ligation dependent probe amplification (MLPA) or chromosomal microarray is now typically used for diagnostic testing of patients with clinical features suggestive of 22q11.2DS. Prior to molecular testing, the most common genetic testing was using fluorescence in situ hybridization (FISH) with probes in the A-B region (**Figure 1**). Therefore, individuals that had larger deletions or non-overlapping deletions were previously known. The most frequent deletion is a 3 million base pair (Mb) hemizygous deletion flanked by LCR22s A-D (A-D). Nested A-B (1.5 Mb) and A-C (1.8 Mb) deletions occur with lower frequency but with similar overall phenotypes (**Figure 1**) [8–11].

**Figure 1.**
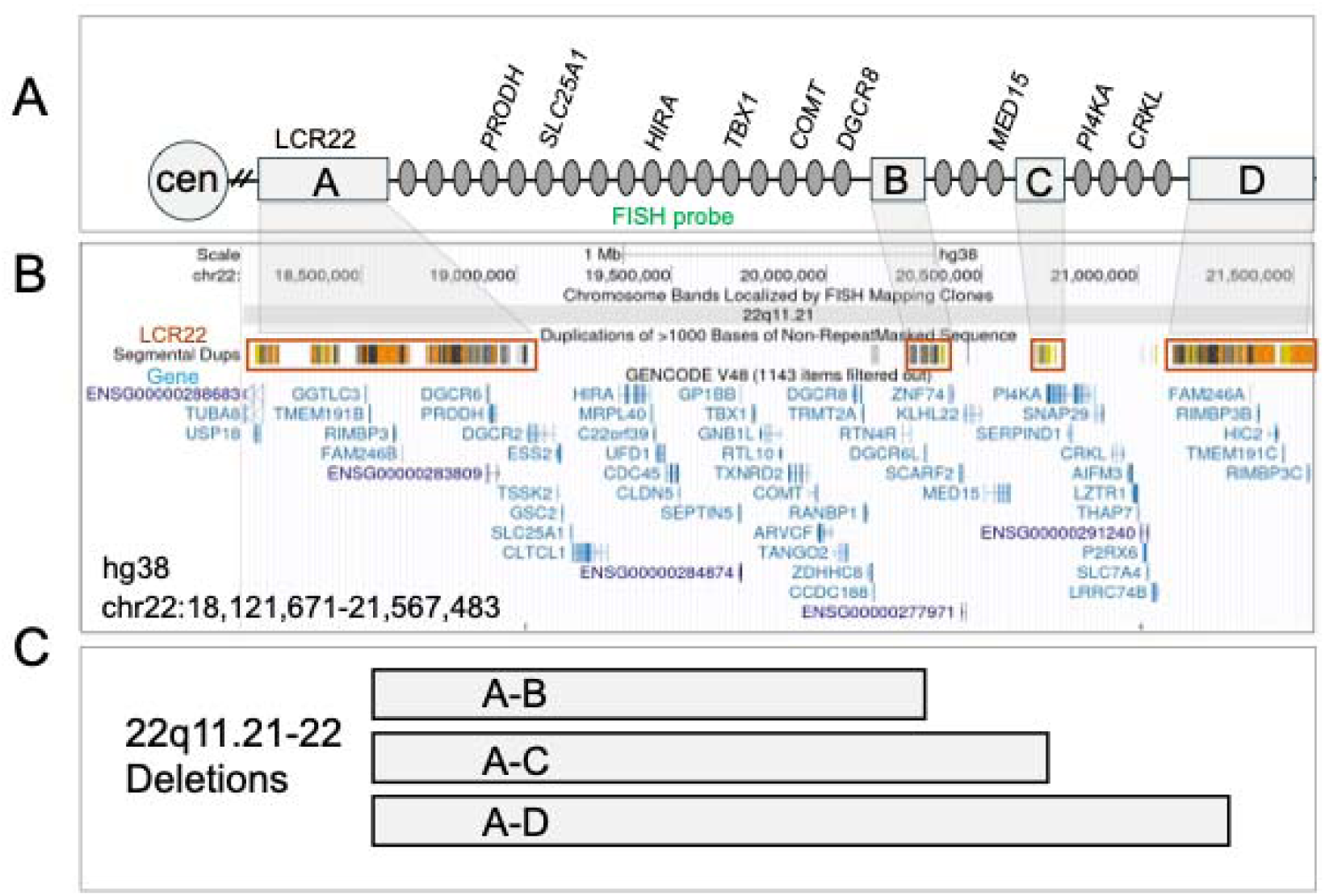
Map of 22q11.2 region. (A) Diagram of the 22q11.21-22 region with the centromere (cen) shown on the left. The position of the LCR22s (low copy repeats on 22q11.2), -A, -B, -C, - D are shown as light gray boxes. Genes are indicated (gray ovals) with representative known coding genes, between the LCR22s. (B) Screenshot from the UCSC browser, hg38 assembly (coordinates are indicated). The position of the LCR22s (segmental duplications are shown (orange boxes) as are the genes spanning the region (Genecode track). (C) The position of the deletions in the 22q11.2 region, A-B, A-C and A-D, correspond to chromosome breakpoints in the respective LCR22s.

The clinical presentation of this syndrome ranges from mild to severe including characteristic facial differences, immune deficits, and hypocalcemia as well as neurocognitive or psychiatric disorders[12 13]. One of the most frequent and serious medical findings is congenital heart disease (CHD) that occurs in 50-80% of individuals with 22q11.2DS; this range in percentage is a result of ascertainment in different clinical settings[14]. The types of CHD that occur range from isolated, mild aortic arch or arterial branching defects (right sided aortic arch, RAA; abnormal origin of the right or left subclavian arteries, ASA) with little clinical symptoms unless accompanied with a vascular ring, ventricular and/or atrial septal defects (septal defects, SD; VSD, ASD) to severe, complex and potentially lethal conotruncal heart defects (CTDs) including tetralogy of Fallot (TOF), interrupted aortic arch type B (IAAB) and persistent truncus arteriosus (PTA)[15 16]. The most severe CTDs require surgery for survival beyond the neonatal or early infancy period. Overall, there is extensive phenotypic heterogeneity in the prevalence and types of CHD in 22q11.2DS. The role of basic features on phenotypic heterogeneity of CHD is not well established due to the rarity of the syndrome and lack of a comprehensive and systematic assessment of this type of medical data.

In this study, we retrospectively analyzed demographic, deletion-size, genome-wide genetic principal component (PC), and detailed postnatal cardiac phenotype data from 3,016 unrelated individuals with 22q11.2DS ascertained at specialized clinical centers in the US, Canada, Europe, South America, Israel, and Australia. The cohort size and harmonized phenotype curation enabled lesion-specific analyses, while the multisite design required multivariable models to account for site-level heterogeneity and correlated covariates. This approach identified a set of deletion-size and genome-wide PC associations with specific CHD phenotypes.

## METHODS

### Ethics approval statement

Our research on 22q11.2 disorders is part of an Internal Review Board approved program at Albert Einstein College of Medicine (IRB #1999-201). Individuals ascertained at Albert Einstein College of Medicine or Montefiore Medical Center obtained written informed consent (and/or assent for minors) for all participants as part of this IRB protocol. Co-authors at different sites have their own individual institutional IRB approval with their own informed consent procedures. This includes, Children’s Hospital of Philadelphia (IRB 07-5352), University of Pennsylvania (IRB #832678), University of California Los Angeles (IRB#10-001071-CR-00012), Medical College of Wisconsin (MCW PRO 47811; v. 2025.07.30), Duke University (IRB Pro00010602), University of Toronto affiliated hospitals (IRB CAMH 114-2001, CAMH 154-2002, CAMH 150-2002, CAMH 151-2002 and UHN 98-E156), Sapienza (Protocol Ref. 5803) and Rome, IT (Prot. 468/12), University of Geneva (IRB Project ID: 2020-02296), Geneva Ethic Committee (Marco Armando). Further, this study was approved by ethics committees in the UK, The Netherlands (Utrecht; 08-345 Vorstman) and Belgium (Cardiff, NHS Wales Research Ethics Committee; Maastricht, Maastricht University Medical Centre-IRB NL70681.068.19; KU Leuven; Research Medical Ethics Committee UZ KU Leuven, IRB S52418), University of Newcastle, Newcastle, AU, Son Espases University Hospital, Mallorca-IRB# IB 4084/20, University Hospital La Paz, Madrid, Royal College of Surgeons in Ireland, Dublin, Ireland, King’s College London, London, Polish Mother’s Memorial Hospital-Research Institute, Tel Aviv University-Sheba Medical Center Israel (IRS#SMC-7783-10). The study was approved by the local ethics committee at the Institute of Psychiatry, South London and Maudsley Trust (067/00; L. Campbell). All individuals were de-identified for this study.

### Study population

De-identified data were evaluated from an original sample of 3,746 unrelated individuals with 22q11.2DS recruited from clinical centers for 22q11.2DS in the US, Canada, Northern and Southern Europe, Chile and Australia, including and in addition to the International 22q11.2 Brain and Behavior Consortium [17] (Full de-identified dataset is presented in **Supplementary Table S1**). The ascertainment varied at different sites and included pediatric genetics clinics, cardiology services, adult psychiatric clinics and other centers of care. Each had a clinical diagnosis of 22q11.2DS (velo-cardio-facial syndrome-MIM#192430, DiGeorge syndrome-MIM#188400, conotruncal anomaly face syndrome). The type of the 22q11.2 deletion was determined using molecular diagnostic testing, including chromosomal microarray, multiplex ligation-dependent probe amplification (MLPA; SALSA MLPA kit P250 DiGeorge; MRC Holland, The Netherlands[18]) or research based whole exome/genome sequencing[19–21]. We included subjects with deletions between LCR22 A-B, A-C and A-D in this study and excluded the rare subjects with nested distal deletions such as B-D and C-D deletions. Subjects with other deletion types on 22q11.2, or those extending centromeric or telomeric to this region, were extremely rare and were also excluded. These B-D, C-D, or rare, atypical deletions were excluded due to the paucity of these subjects in our cohort.

Most individuals with 22q11.2DS were unrelated probands and had a *de novo* deletion. We had family pedigree data from the immediate family members for most subjects. Transmitting parents and relatives with 22q11.2DS were excluded for consistency. A total of 3,016 subjects fulfilled the study criteria and had cardiology and echocardiography reports from infancy or childhood. Children were included when the clinical cardiac record documented either a lesion at birth or a normal heart; adults were included only when pediatric cardiac evaluation records, including echocardiography, were available. Prenatal-only cases were excluded because fetal ascertainment is enriched for severe or ultrasound-detectable lesions, pregnancy outcomes and postnatal echocardiographic confirmation were not uniformly available across sites, and inclusion would have introduced a different ascertainment frame from the postnatal clinical cohort. Prenatal incidence is an important complementary question but requires a separate population-based fetal analysis. We obtained sex, self-reported race and/or ethnicity (race/ethnicity), and recruitment-site information for most subjects. Self-reported information was obtained from the proband, parent, or guardian. There was missing data on sex in less than 0.5% and self-reported race/ethnicity in about 13%. A portion of these data in a subset of the 3,016 subjects was previously described[19–21].

### Association between sex or self-reported race/ethnicity and deletion type

Whole genome sequence (WGS) was available for 1,868 of the 3,016 individuals (61.9%) included in the study. Exclusion of first-degree kinship among these 1,868 subjects was confirmed using KING software[22]. For subjects with WGS, we used genetically inferred sex and excluded sex mismatches before analysis; for the 1,148 without WGS data, we used patient-identified sex. Proband sex (male, female) was compared across the three 22q11.2 deletion groups (A-B, A-C, and A-D) using a 2 x 3 contingency table and Pearson chi-square test. Self-reported race/ethnicity was defined as White, Black or African American, Asian, American Indian or Alaska Native, Native Hawaiian or Other Pacific Islander, Mixed race/ethnicity, Other, or missing. Mixed race/ethnicity was defined as more than one self-reported category. Because smaller groups were sparse, deletion-type analyses using self-reported race/ethnicity were restricted to the two largest categories: White and Mixed. Statistical significance was defined as adjusted P < 0.05.

### Co-occurrence of cardiac phenotypes in 22q11.2DS

Cardiac lesions were organized into broad classes and phenotype subgroups to parse individual and complex lesions while counting each subject once for each modeled outcome. Broad classes included any CHD, conotruncal defects (CTD), and isolated septal defects (SD; ASD-only, VSD-only, or ASD+VSD without CTD or aortic arch anomalies). Phenotype subgroups, defined in Supplementary Table S2, included TOF, PTA, IAAB, VSD-only, ASD-only, ASD+VSD, RAA+, RAA-only, PS/PA+, PS/PA-only, and ASA. For co-occurrence analyses, we generated 2 x 2 contingency tables for all phenotype pairs with non-missing data and testable marginal totals, estimated odds ratios using two-sided Fisher exact tests, and controlled false discovery with the Benjamini-Hochberg procedure. Pairwise relationships were visualized with a signed score that combined the direction of association with the FDR-adjusted q value, followed by hierarchical clustering using pheatmap.

### Principal component analysis following removal of batch effects in whole genotype sequence

We derived the top five principal components (PCs) from common autosomal single nucleotide variants in the 1,868 subjects with available short-read WGS. These samples came from two publicly available sequencing resources: the International 22q11.2 Brain and Behavior Consortium/NIMH Data Archive and dbGaP/CIDR (phs002514.v1.p1). Variants were processed with the DRAGEN unified variant-calling pipeline[23], then filtered for depth, genotype quality, missingness, variant quality, and linkage disequilibrium before PCA. This QC step was performed to minimize platform-related batch effects; after QC, none of the top five PCs used in the association analysis was associated with WGS platform status (**Supplementary Figure S1**).

### Ancestry analysis

Global ancestry proportions for subjects with WGS were estimated using ADMIXTURE v1.3.0[24]. After standard PLINK quality-control filters for missingness, minor allele frequency, Hardy-Weinberg equilibrium, heterozygosity, and linkage disequilibrium, the cohort was merged with 1000 Genomes reference samples[25]. ADMIXTURE was run with k=5 to estimate continental ancestry components (Admixed American [AMR], East Asian [EAS], South Asian [SAS], European [EUR], and African [AFR]), which were visualized with pong v1.5.

### Univariable screening

Each CHD class and phenotype subgroup was first explored in relation to one explanatory variable at a time. Categorical variables (recruitment site, sex, self-reported race/ethnicity, and deletion type) were tested using contingency-table analyses and crude logistic-regression models. For PC analyses, each phenotype was regressed on the five Z-standardized PC scores, and a likelihood-ratio test compared the full PC model with an intercept-only model. False discovery was controlled using the Benjamini-Hochberg procedure with q < 0.05 considered statistically significant.

### Multivariable mixed-effects logistic regression

For each binary phenotype group, we fitted a separate mixed-effects logistic-regression model with 22q11.2 deletion category (reference: A-B), sex, and PC1-PC5 as fixed effects. Recruitment site was modeled as a random intercept to account for center-level differences in diagnosis and ascertainment, stabilize estimates for sites with small sample sizes, and avoid overfitting that would arise if each site were treated as a fixed covariate.

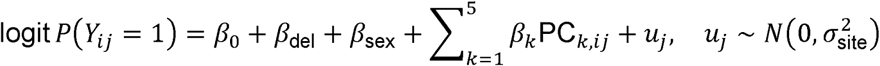

Models were fitted with lme4::glmer (optimizer = bobyqa) in R.

Wald z-tests provided a two-sided P value for every non-intercept coefficient. We controlled the false discovery rate (FDR) within each phenotype (column q in Supplementary Tables) and then applied a second Benjamini-Hochberg adjustment across the complete set of 112 non-intercept fixed-effect estimates (14 phenotypes x 8 predictors), reported as q_global. Results with q_global < 0.05 were considered study-wide significant.

### Bootstrap sensitivity analysis

To assess the robustness of key associations identified in the mixed-effects logistic regression models, we performed a two-stage nonparametric bootstrap procedure that preserved the hierarchical structure of the data. First, recruitment sites were resampled with replacement.

Second, within each resampled site, individuals were resampled with replacement. For each bootstrap replicate, we refit the same mixed-effects logistic regression model used in the primary analysis, including deletion group, sex, and ancestry principal components as fixed effects and study site as a random intercept. We repeated this procedure for 500 iterations for selected significant associations (PS/PA+ with PC2, PTA and SD with A-D deletion, and ASA with PC4). From the bootstrap distributions of the fixed-effect coefficients, we estimated median effects, empirical 95% confidence intervals, and the proportion of replicates with effect estimates in the same direction as the original model.

## RESULTS

### Demographic features in 3,016 subjects with 22q11.2DS

We analyzed 3,016 unrelated postnatal subjects with 22q11.2DS. Cohort characteristics and cardiac phenotype prevalences are summarized in **Table 1**. The common A-D deletion was present in 2,788 individuals (92.4%), while 172 (5.7%) had A-B and 56 (1.9%) had A-C deletions. Sex was balanced (1,493 male, 49.5%; 1,523 female, 50.5%). Self-reported race/ethnicity was most often White (2,034, 67.4%) or Mixed (379, 12.6%); 401 individuals (13.3%) were missing race/ethnicity. Recruitment-site information was available for all but one subject across 22 recorded site categories, with the largest contributions from Philadelphia, Bronx, and Toronto. WGS data are available for 1,868 individuals (61.9%).

**Table 1.** Cohort characteristics and cardiac phenotype prevalence.

| *Category* | *Measure* | *n (%)* |
| --- | --- | --- |
| Cohort | Total analyzed | 3,016 |
| Deletion type | A-D | 2,788 (92.4%) |
|  | A-B | 172 (5.7%) |
|  | A-C | 56 (1.9%) |
| Sex | Male | 1,493 (49.5%) |
|  | Female | 1,523 (50.5%) |
| Self-reported race/ethnicity | White | 2,034 (67.4%) |
|  | Mixed | 379 (12.6%) |
|  | Other (Black, Asian, American Indian/Alaska Native, Native Hawaiian/Other Pacific Islander) | 202 (6.7%) |
|  | Missing | 401 (13.3%) |
| Genomic data | Whole-genome sequencing / PCA | 1,868 (61.9%) |
|  | Non-missing ADMIXTURE proportions | 1,785 (59.2%) |
| CHD class | Any CHD | 1,793 (59.4%) |
|  | Conotruncal defect (CTD) | 1,193 (39.6%) |
|  | Septal defect (SD) | 424 (14.1%) |
| CHD subgroup | Tetralogy of Fallot (TOF) | 599 (19.9%) |
|  | Right-sided aortic arch with/without other defects (RAA+) | 364 (12.1%) |
|  | Isolated ventricular septal defect (VSD-only) | 253 (8.4%) |
|  | Interrupted aortic arch type B (IAAB) | 214 (7.1%) |
|  | Persistent truncus arteriosus (PTA) | 131 (4.3%) |
|  | Abnormal origin of the subclavian arteries (ASA) | 119 (3.9%) |
|  | Pulmonary stenosis/atresia with other lesions excluding TOF (PS/PA+) | 107 (3.5%) |
|  | Isolated atrial septal defect (ASD-only) | 92 (3.1%) |
|  | ASD with VSD but no other defects (ASD+VSD) | 79 (2.6%) |
|  | Pulmonary stenosis/atresia without other defects (PS/PA-only) | 22 (0.7%) |

### Association analysis between deletion type with sex and self-reported race/ethnicity

For the first analysis, reported sex (including genetically validated sex from WGS) was compared across the A-B, A-C, and A-D deletion types. The proportion of males was similar across deletion groups (**Supplementary Table S3**), and Pearson chi-square testing confirmed that sex distribution did not vary by deletion type (chi-square = 1.0, df = 2, P = 0.61). We then tested whether the deletion type differed between the two largest self-reported race/ethnicity groups, White and Mixed. The distribution was similar between these two groups (**Supplementary Table S4**; chi-square = 2.3, df = 2, P = 0.32). Thus, deletion type was not associated with sex or, within the two largest self-reported race/ethnicity strata, with White versus Mixed race/ethnicity. The study was not powered to test deletion-type differences across the smaller self-reported race/ethnicity groups.

### Ancestry in the 22q11.2DS cohort from WGS

To more precisely identify genetic ancestry in the 22q11.2DS population, we used 1000 Genomes reference data[25] and ADMIXTURE to estimate global ancestry proportions in subjects with WGS. Among the 1,785 subjects with non-missing ADMIXTURE estimates, the mean ancestry proportions were predominantly European (EUR 81.5%), with smaller mean proportions of African (AFR 5.6%), Admixed American (AMR 5.2%), South Asian (SAS 5.2%), and East Asian (EAS 2.5%) ancestry. These estimates confirmed that the WGS subset was majority European ancestry while retaining measurable diversity and admixture as shown in Figure 2. We used these proportions below as sensitivity variables to assess whether PC-associated CHD findings were explained by direct continental ancestry estimates.

**Figure 2.**
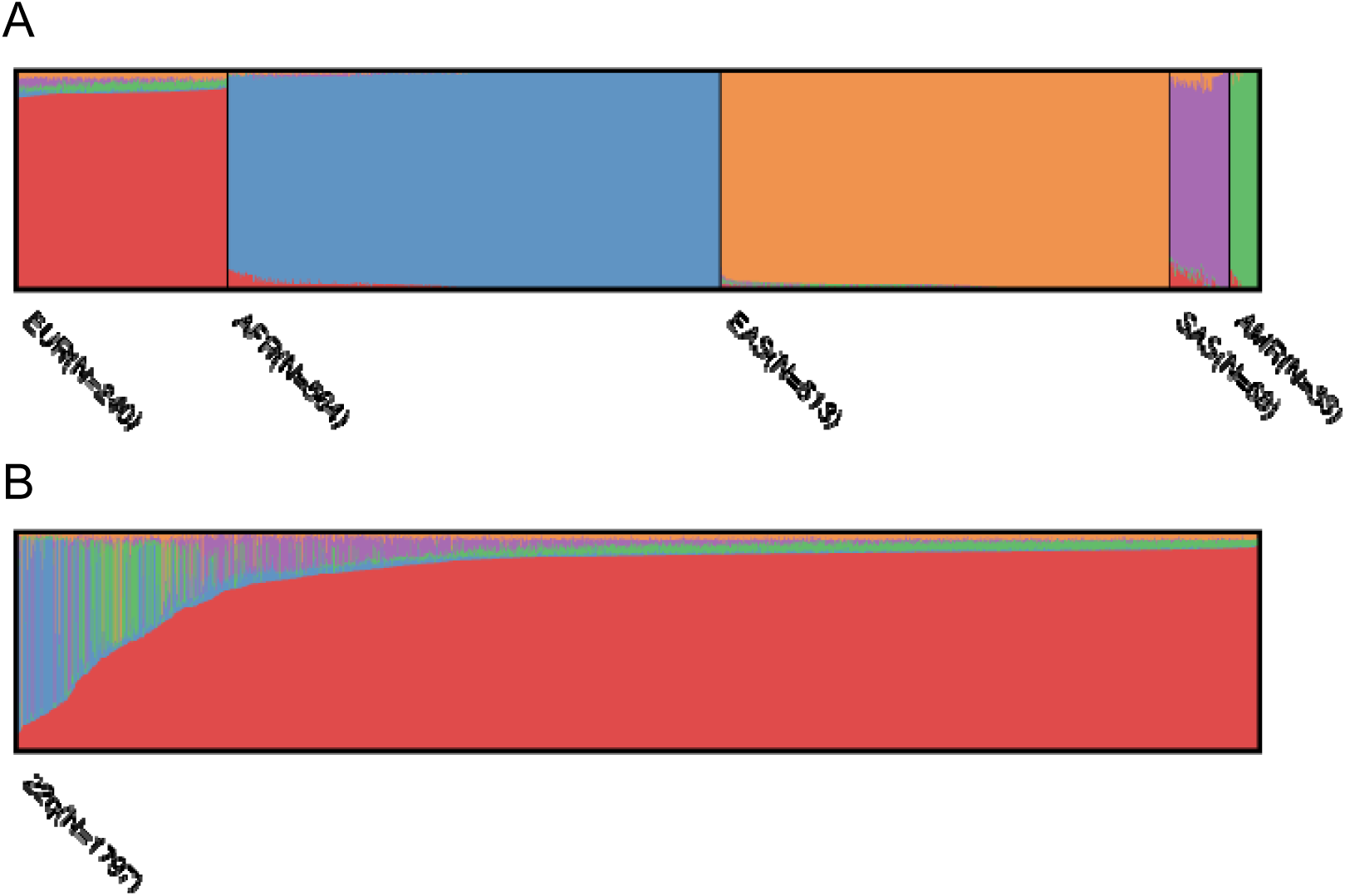
Analysis of 22q population structure. (A) Admixture plot (k=5) for 1000 Genomes reference individuals, defining the global ancestry color mapping. (B) Admixture plot (k=5) all 1,797 cohort individuals, where each bar represents the estimated global ancestry of an individual. Colors correspond to predominant global ancestry as defined in (A): red represents European ancestry (EUR), purple represents South Asian ancestry (SAS), blue represents African ancestry (AFR), green represents Admixed American (AMR), and orange represents East Asian ancestry (EAS).

### Architecture of CHD types in 22q11.2DS

In the cohort, 1,793 individuals (59.4%) had CHD (**Table 1**). CTDs were present in 1,193 individuals (39.6% of total; 66.5% of CHD), and isolated septal defects were present in 424 (14.1% of total; 23.6% of CHD). Remaining CHD phenotypes included isolated aortic arch/arterial branching or pulmonary stenosis/atresia lesions not assigned to CTD or SD. We generated 11 phenotype subgroups (TOF, RAA+, VSD-only, IAAB, RAA-only, PTA, ASA, PS/PA+, ASD-only, ASD+VSD, and PS/PA-only; **Supplementary Table S2**). Pairwise co-occurrence analyses (**Supplementary Table S5**) confirmed expected mutual exclusivity among several major diagnoses and enrichment of RAA with TOF (OR = 2.52, q = 5.24E-13) and PTA (OR = 1.84, q = 1.35E-2), consistent with the known enrichment of RAA among CTDs in 22q11.2DS[15 26]. These classes and subgroups were used for the association analyses.

### Deletion size and genetic background contribute to heterogeneity in CHD risk

Preliminary univariable screening analyses indicated that the prevalence of CHD classes and phenotype subgroups varied across recruitment sites and showed associations with sex, deletion size, and genetic PCs for a subset of cardiac lesions (**Supplementary Tables S6-S10**). The observed site-to-site heterogeneity likely reflects differences in referral pathways, including pediatric cardiology, genetics, adult psychiatry, and specialty 22q11.2DS clinics; local echocardiography documentation practices; age at enrollment; survival of severe neonatal lesions; and regional genetic-testing patterns. These observations motivated the use of a multivariable mixed-effects framework to jointly model covariates while accounting for recruitment-site heterogeneity.

We applied multivariable mixed-effects logistic regression to model each CHD class and phenotype subgroup as the outcome while adjusting for deletion size, sex, and PC1-PC5, with recruitment site included as a random intercept in the statistical analysis. This framework accounted for measured covariates and unmeasured site-level differences, such as diagnostic practices and ascertainment patterns, and provided estimates of independent associations in the combined cohort.

Across 14 CHD outcomes and 112 non-intercept fixed-effect contrasts, four associations remained significant after study-wide FDR control (q_global < 0.05; **Table 2**, **Supplementary Table S11**). These involved a subset of CHD phenotypes associated with 22q11.2DS.

**Table 2.** Significant fixed-effect coefficients in the multivariable mixed-effects logistic regression.

| *CHD* | *Predictor* | *OR (95% CI)* | *P-value* | *q<sub>global</sub>* |
| --- | --- | --- | --- | --- |
| PTA | Deletion A-D vs A-B | 0.37 (0.18 - 0.75) | 5.74E-3 | 4.52E-2 |
| SD | Deletion A-D vs A-B | 4.68 (1.71 - 12.84) | 2.70E-3 | 2.43E-2 |
| PS/PA+ | PC2 | 0.73 (0.61 - 0.87) | 5.35E-4 | 5.19E-3 |
| ASA | PC4 | 2.59 (1.37 - 4.89) | 3.37E-3 | 2.83E-2 |

Notes: The statistical significance for these regression analyses was set at the global FDR threshold ( $q_{\text{global}} < 0.05$ ). PTA, persistent truncus arteriosus; SD, septal defect including atrial or ventricular septal defect; PS/PA+, pulmonary stenosis or atresia with other heart lesions excluding TOF; ASA, abnormal origin of the subclavian arteries. PC, principal component. OR, odds ratio.

Deletion size showed significant associations with PTA and SD, but these estimates should be interpreted cautiously because A-B deletions were much less frequent than A-D deletions (172 versus 2,788 individuals). Compared with individuals with the smaller A-B deletion, individuals with the A-D deletion had lower odds of PTA (**Table 2**; OR = 0.37, 95% CI: 0.18-0.75, q_global = 0.045). Conversely, the A-D deletion was associated with higher odds of SD (**Table 2**; OR = 4.68, 95% CI: 1.71-12.83, q_global = 0.024). In two-stage bootstrap sensitivity analyses that resampled recruitment sites and individuals within sites, the direction of effect for the A-D deletion was preserved for both PTA (median OR = 0.37; P[OR < 1] = 0.90) and SD (median OR = 5.2; P[OR > 1] = 0.99). This analysis evaluates the stability of the findings while accounting for clustering by recruitment site, and would in the future benefit from replication studies in independent cohorts.

We also observed significant associations between two CHD subtypes and genetic PCs. PS/PA+ was associated with a 27% reduction in odds for every one-standard-deviation increase along PC2 (OR = 0.73, 95% CI: 0.61-0.87, q_global = 0.0052), with bootstrap analyses supporting the direction of this effect in approximately 92% of bootstrap samples. ASA was associated with PC4 (OR = 2.59, 95% CI: 1.37-4.89, q_global = 0.028). Bootstrap analyses showed a median OR of approximately 3.1, with approximately 96% of bootstrap samples yielding OR > 1, although confidence intervals were wide. In sensitivity models replacing PCs with ADMIXTURE-derived ancestry proportions, no ancestry-proportion term was significant for PS/PA+ (P = 0.17-0.94) or ASA (P = 0.21-0.59), suggesting that these PC associations were not explained by direct continental ancestry estimates alone.

## DISCUSSION

In this study of 3,016 individuals with 22q11.2DS, we identified four lesion-specific associations that remained significant after multivariable adjustment and study-wide FDR control. Two involved deletion size, and two involved genome-wide PCs. A major strength of this study is the large size of the cohort with available basic medical data. One limitation is that the A-B group was much smaller than the A-D group and another is that PCs are latent axes of genome-wide variation rather than directly interpretable biological mechanisms. The results therefore support a more focused conclusion that the deletion interval and broader genetic context may contribute to CHD variability in 22q11.2DS. Although not currently clinically actionable, this data serves to generate hypotheses that can be examined in additional cohorts.

The deletion-size findings suggest that PTA and SD may not respond uniformly to the same deleted interval in human subjects. The A-D carriers had lower odds of PTA and higher odds of SD than A-B carriers after adjustment for covariates. Prior clinical reviews, systematic reviews, and smaller genotype-phenotype analyses have emphasized high lesion heterogeneity in 22q11.2DS and have not consistently identified this PTA/SD pattern[14 15 27-29]. Thus, our deletion-size estimates should be interpreted as cohort-specific signals that are interesting and that require replication. A plausible biological interpretation is that dosage-sensitive genes in the A-B interval, including TBX1-related pathways, interact with genes or regulatory elements in the B-D interval to influence cardiac progenitor cells in developmental processes[19 30]. However, the confidence intervals remain wide, and the imbalance in deletion-group sizes limits precision. Nevertheless, mixed-effects logistic regression appropriately accommodates unequal group sizes, and bootstrap resampling supported the stability of the observed directions of effect across resampled datasets.

The PC-associated findings are interesting but also require caution in interpretation. PCs derived from common genetic variation are useful for modeling population structure and related genome-wide variation[31], but they do not identify causal alleles or pathways. In this cohort, PC2 correlated most strongly with AMR, EAS, and AFR ancestry proportions, whereas PC4 correlated most strongly with SAS and AMR proportions (**Supplementary Table S12**).

Nevertheless, direct ADMIXTURE-derived ancestry proportions were not independently associated with PS/PA+ or ASA in sensitivity models, suggesting that the PC signals are not simple continental-ancestry effects. One possible explanation is that PCs capture fine-scale ancestry, local haplotypes, regulatory background, or other unmeasured factors that are not well represented by broad ancestry proportions. Long-range chromatin interactions within and beyond the 22q11.2 region have been proposed as one mechanism by which genomic background could influence phenotypic variability[32], although such mechanisms were not directly tested here. Similar modifier effects have been described in broader CHD genetics, where common variation can influence risk even when a primary lesion or rare variant is present[33 34].

Ascertainment remains an important consideration. Severe conotruncal defects such as PTA are associated with substantial early morbidity and mortality, particularly without timely surgical intervention, and individuals with the most severe phenotypes may be underrepresented in postnatal clinical cohorts. Conversely, individuals with milder isolated lesions may be less likely to undergo genetic testing if syndromic features are subtle. Site-level referral patterns may further enrich different centers for pediatric cardiology, genetics, adult psychiatry, or multidisciplinary 22q11.2DS populations. These factors likely contribute to the observed recruitment-site heterogeneity and should be considered when interpreting genotype-phenotype associations.

CHD, including CHD in syndromic contexts, is multifactorial. Maternal diabetes, other metabolic conditions, medication exposures, and additional noninherited factors are associated with CHD risk in the general population[35], but these variables were not available uniformly across the international cohort. Similarly, the current analysis focused on cardiac phenotypes and did not test whether extracardiac manifestations vary by deletion type or genetic background. Several limitations should therefore be considered: the marked imbalance between deletion groups, incomplete WGS availability, potential site-specific ascertainment and documentation differences, lack of uniformly collected prenatal and environmental exposure data, and the limited biological interpretability of PCs. Future analyses that integrate extracardiac phenotypes, prenatal ascertainment, maternal metabolic and environmental exposures, rare variants, polygenic scores, and functional regulatory annotations will be needed to move from statistical association toward clinically useful prediction.

In summary, this study identifies lesion-specific associations between deletion size, genome-wide PCs, and selected CHD phenotypes in a large postnatal 22q11.2DS cohort. The deletion-size findings implicate distinct susceptibility patterns for PTA and SD, whereas the PC findings suggest that broader genetic background may modify risk for PS/PA+ and ASA. Because these associations are modest and not yet mechanistically resolved, they should be viewed as a framework for replication and for future studies of genetic and environmental modifiers of CHD expression in 22q11.2DS.

## Data Availability

Subject demographic data is provided in the supplementary tables in the manuscript. Whole genome sequence (WGS) is available at the International Brain and Behavior Consortium NIMH Data Archive NDA64. Additional WGS is available at dbGAP. NIH Center for Inherited Disease Research phs002514.v1.p1.

https://nda.nih.gov/

https://cidr.jhmi.edu/

## ACKNOWLEDGEMENTS

We warmly thank the families who participated in the study, and the family associations (Generation 22, Connect 22, and Relais 22) for their support. This work was supported by NIH grants P01HD070454, R01HL157157, R01GM125757 and U01MH101720 (BEM, ZDZ, REG, DMM, BSE, PIs). This work was also supported by R21HD060309 (AOM, MEM). This work was supported in part by the Canadian Institutes of Health Research (CIHR) [MOP-313331 and MOP-111238] and from the Inaugural Dalglish Chair in 22q11.2 Deletion Syndrome at the University of Toronto and University Health Network (ASB). The work was supported by ANID-Chile Fondecyt grants #1100131, 1130392, 1171014, 1211411 to GR. Grants from the Swiss National Science Foundation (#320030_179404 and #320030_144260 to SE and MS), The National Center of Competence in Research (NCCR) “Synapsy - The Synaptic Bases of Mental Diseases” (#51NF40-185897 to SE) and NeuroNA-Synapsy Clinical Cohort Grant (to SE and MS). Grants U01MH119736, R01 MH085953 and SFARI Explorer Award provided support (CB); as well as from KU Leuven (C14/24/135) and FWO grants G0A2622N and G0A7T26N (JRV, JB, AS). JASV is the holder of the SickKids Psychiatry Associates Chair in Developmental Psychopathology and acknowledges funding support (U01MH101722).

## DATA AVAILABILITY

Subject demographic data is provided in the supplementary tables in the manuscript. WGS is available at the International Brain and Behavior Consortium-NIMH Data Archive-NDA64. Additional WGS is available at dbGAP; NIH Center for Inherited Disease Research; phs002514.v1.p1).

## SUPPLEMENTARY TABLE LEGENDS

**Supplementary Table S1. Demographic and CHD phenotype data for 22q11.2DS cohort of 3,016 total subjects.** Column A is the record identification of each subject. Column B is the study site listed by city, state and country. Column C is the sex of each subject (1 is male and 2 is female). Column D is self-reported ancestry (0 is unknown, 1 is American Indian or Alaska Native, 2 is Asian, 3 is Black or African American, 4 is Native Hawaiian or Other Pacific Islander, 5 is White, 6 is mixed of different ancestries, 7 is other ancestry not fitting into the other categories). Column E is the deletion size (A-D, A-B, or A-C). Columns F-J are the principal component results for each subject. Columns K-X are phenotypes in which 0 means unaffected with that phenotype and 1 means affected with that phenotype, while NA means there is missing data for that defect. CHD is any heart defect, CTD is any conotruncal defect minus SDs (ASD-only, VSD-only and ASD+VSD). Phenotype subgroups are listed in separate columns (see definitions in Supplementary Table S2; TOF, SD, RAA+ that is RAA with/out other defects, VSD-only is isolated VSD, IAAB, RAA-only that is RAA without other defects, PTA, ASA without other defects, PS/PA+ is PS or PA not part of TOF but can be with other defects, ASD-only is isolated ASD, ASD+VSD is ASD with VSD but no other defects, PS/PA-only is PS or PA but no other defects). Columns Y-AC are genetic admixture ancestry information for each subject; AMR is Admixed American, EAS is East Asian, SAS is South Asian, EUR is European and AFR is African.

**Supplementary Table S2. Formatted definitions of cardiac phenotypes observed in the 22q11.2DS cohort.** The table lists each phenotype variable, abbreviation, grouping class, and operational definition used for the mutually exclusive CHD classes and phenotype subgroups.

**Supplementary Table S3. There is no difference in the percentage of sex among 22q11.2DS subjects with different deletion types.** Sex versus deletion types were compared and percentages are provided.

**Supplementary Table S4. There is no difference between self-reported White versus mixed ancestry and deletion type in the 22q11.2DS cohort.** Self-reported ancestry of the two largest groups, of White and mixed ancestry is provided with deletion type. There were no significant differences in deletion type.

**Supplementary Table S5. Mutually exclusive CHD phenotypes in the 22q11.2DS cohort.** Each phenotype was compared to each other (Columns A and B) and examined in the total number of pairs possible (Column C) and tested if they co-occur. The odds ratio (OR; Column E) of their co-occurrence is shown along with the p-value (Column F) and q value (Column G) for the cohort.

**Supplementary Table S6. Univariable logistic regression analysis for study site.** Each CHD phenotype (Column A) was compared to the recruitment study site location (Column B). The odds ratio (OR; Column C), lower and upper confidence limits (lcl, ucl; Columns D and E), p-value (Column F), chi-square p value (chisq_p; Column G), and q values for the p-value adjusted for false positives (Column H) are indicated. Significant q values are shown in red font in Column H.

**Supplementary Table S7. Univariable logistic regression analysis for sex.** Each CHD phenotype (Column A) was compared to subject sex (Column B). The odds ratio (OR; Column C), lower and upper confidence limits (lcl, ucl; Columns D and E), p-value (Column F), chi-square p value (chisq_p; Column G), and q values for the p-value adjusted for false positives (Column H) are indicated. Significant q values are shown in red font in Column H.

**Supplementary Table S8. Univariable logistic regression analysis for self-reported ancestry.** Each CHD phenotype (Column A) was compared to self-reported ancestry (Column B), coded as 2 is Asian, 3 is Black or African American, 4 is Native Hawaiian or Other Pacific Islander, 5 is White, 6 is mixed of different ancestries, and 7 is other ancestry not fitting into the other categories. The odds ratio (OR; Column C), lower and upper confidence limits (lcl, ucl; Columns D and E), p-value (Column F), chi-square p value (chisq_p; Column G), and q values for the p-value adjusted for false positives (Column H) are indicated. There are no significant q values.

**Supplementary Table S9. Univariable logistic regression analysis for deletion type.** Each CHD phenotype (Column A) was compared to deletion type (Column B). The odds ratio (OR; Column C), lower and upper confidence limits (lcl, ucl; Columns D and E), p-value (Column F), chi-square p value (chisq_p; Column G), and q values for the p-value adjusted for false positives (Column H) are indicated. Significant q values are shown in red font in Column H.

**Supplementary Table S10. Univariable logistic regression analysis for principal components.** Each CHD phenotype (Column A) was compared to the PC number (Column B). The odds ratio (OR; Column C), lower and upper confidence limits (lcl, ucl; Columns D and E), p-value (Column F), and q values for the p-value adjusted for false positives (Column G) are indicated. Significant q values are shown in red font in Column G.

**Supplementary Table S11. Multivariable mixed-effects logistic regression results for CHD phenotypes.** Each congenital heart disease phenotype is listed in Column A. Column B indicates the model effect type (fixed effect), and Column C specifies the predictor term. Columns D and E report the regression coefficient (Estimate) and its standard error (STD.error) on the log-odds scale. The corresponding odds ratio (OR) is shown in Column F, with the 95% confidence interval given in Columns G-H. Column I reports the Wald z-statistic, and Column J provides the two-sided p-value. Column K shows the false discovery rate (FDR)-adjusted p-value within phenotype, while Column L reports the study-wide FDR-adjusted q-value across all non-intercept phenotype-predictor tests. The recruitment site was modeled as a random effect in all analyses. Non-intercept associations meeting study-wide FDR significance are highlighted.

**Supplementary Table S12. Correlation between principal components and ancestry proportions.** Pearson correlation coefficients (r) and corresponding two-sided p-values are shown for the association between the first five principal components (PC1-PC5; rows) and ADMIXTURE-derived ancestry proportions (columns: East Asian [EAS], South Asian [SAS], European [EUR], African [AFR], and Admixed American [AMR]). Correlations were computed using individuals with non-missing values for both variables. This table is used to aid the interpretation of ancestry-related principal components identified in multivariable analyses.

