## Supplemental Figure S1 for "Deletion size and background genetic variation shape congenital heart disease phenotypes in 3,016 individuals with 22q11.2 deletion syndrome"

Supplementary Figure S1

| A  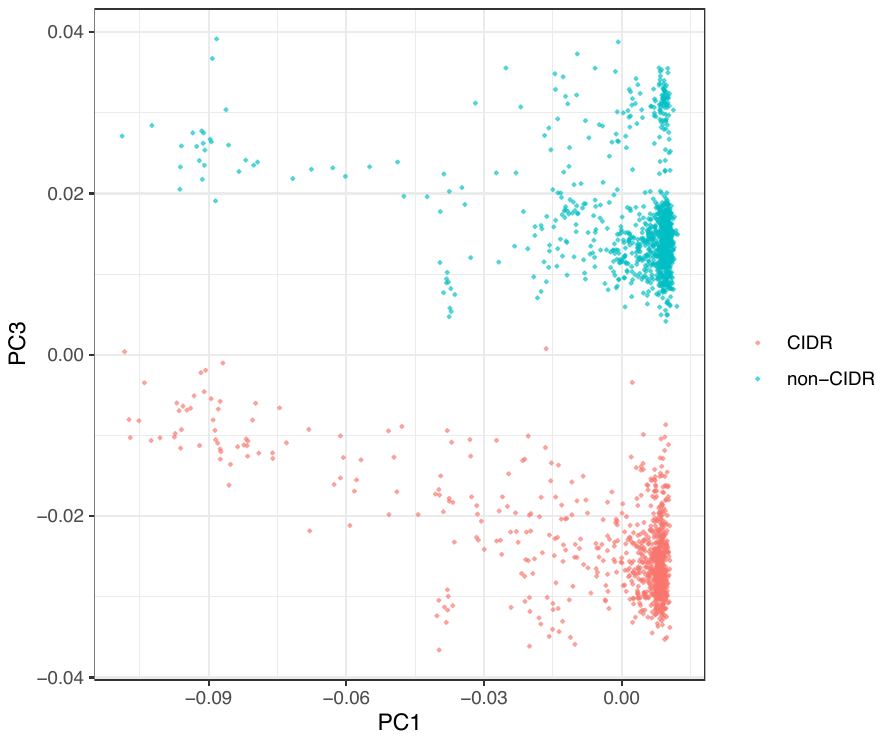 | B  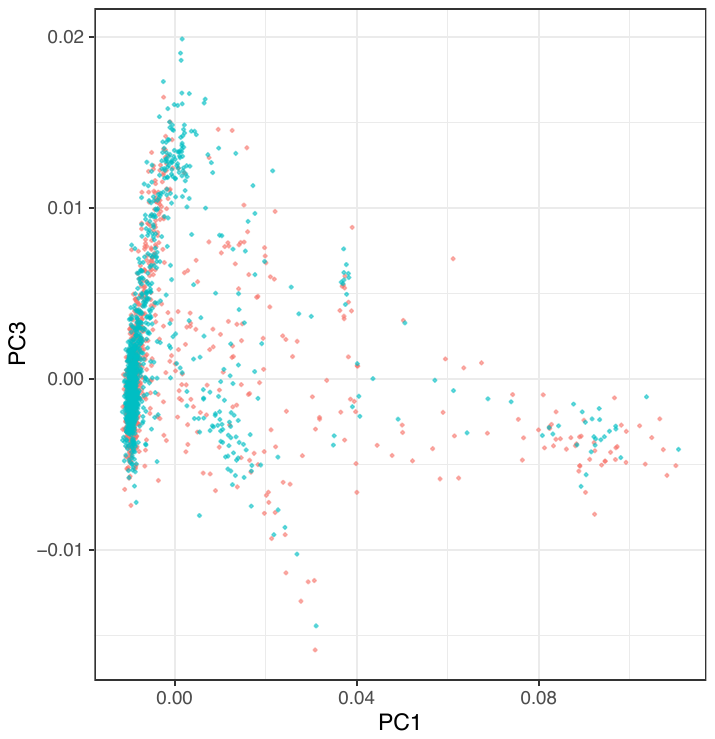 |
| --- | --- |
| C  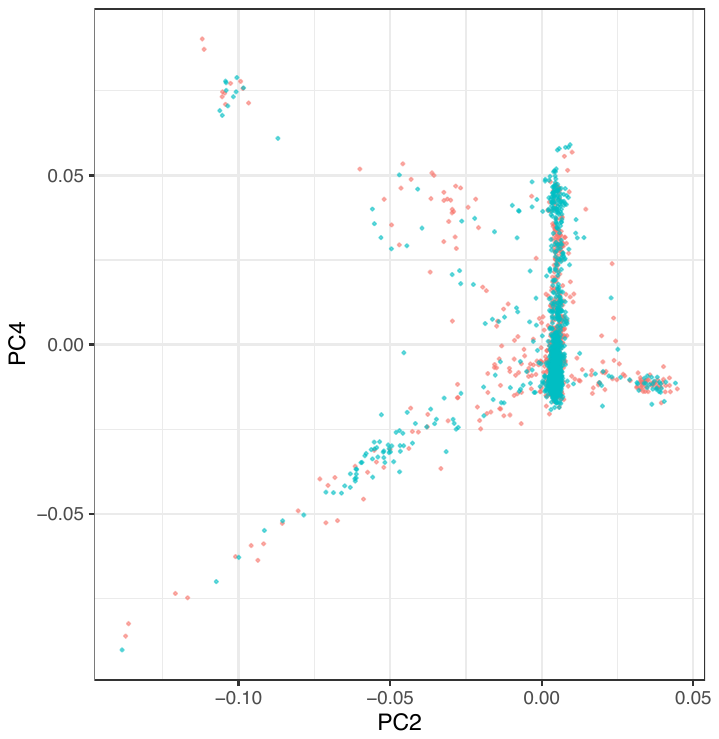 | D  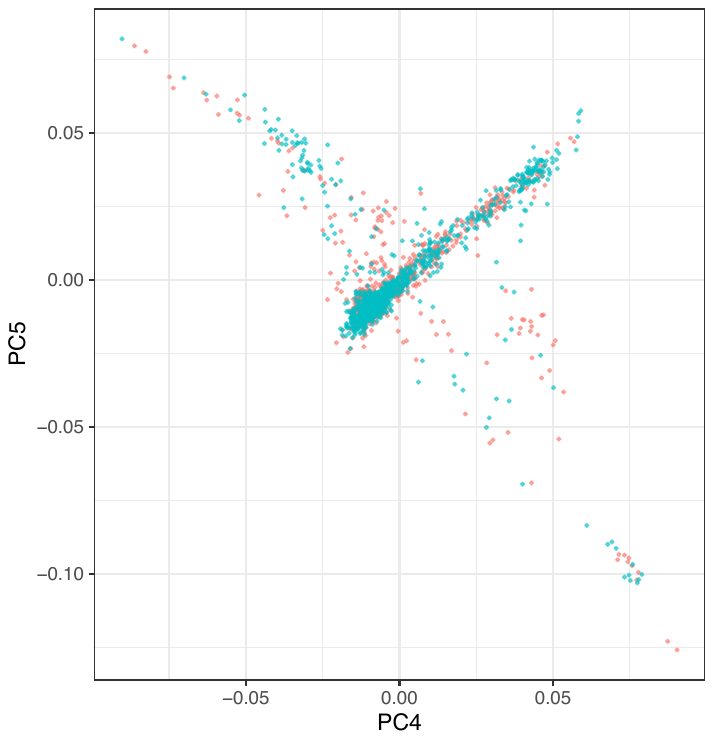 |

**Supplementary Figure S1. Principal-component scatterplot of cohort subjects colored by two batches of WGS data.** (A) PC1-PC3 derived from raw WGS data. B–D, The PCA results derived from WGS data after QC for PC1-PC3 (B), PC2-PC4 (C), and PC4-PC5 (D).

We derived the top five principal components (PCs) from principle component analysis (PCA) to capture variance in population structure based on common variants (MAF < 0.05) from 1,868 subjects with available whole genome sequencing (WGS) data. Those 1,881 subjects include both CIDR and non-CIDR subjects, who were processed using the unified variant calling pipeline but sequenced on different platforms. These platform differences can introduce batch effects, which could be captured by the top PCs (**Supplementary Figure S1A**). To resolve this issue, we performed a variant quality control process (QC) before PCA aimed at removing the observed batch effect. Specifically, we set genotypes with depth (DP) < 5 or genotype quality (GQ) < 10 to missing in the project-level VCF (pVCF) for the WGS data. We then excluded variants with a missingness rate > 10%, in addition to variants with a QUAL score < 30. Finally, we used PLINK to select 300,067 linkage disequilibrium (LD)-pruned common variants on the autosomes with “indep-pairwise” function (window size = 50 SNPs, step size = 10, *r*² threshold = 0.1) and performed PCA. After QC, the batch effect previously observed in PC3 was no longer present, and none of the top five PCs used in our analysis were associated with CIDR status (**Supplementary Figure S1B, S1C, and S1D**).
